# Machine Learning for Predicting Thrombotic Recurrence in Antiphospholipid Syndrome

**DOI:** 10.1101/2025.03.11.25323564

**Authors:** Ana Marco-Rico, Ihosvany Fernández-Bello, Jorge Mateo-Sotos, Pascual Marco-Vera

## Abstract

Thrombotic Antiphospholipid Syndrome (TAPS) is an autoimmune disorder associated with a high risk of recurrent thromboembolic events. Despite advances in anticoagulation, predicting recurrence remains challenging, underscoring the need for more precise risk stratification to optimize personalized treatment. Traditional predictive models struggle to integrate the complexity of clinical and biochemical risk factors, creating an opportunity for Machine Learning (ML) to enhance prognostic accuracy. In this study, we evaluated the performance of the Extreme Gradient Boosting (XGBoost) model in predicting recurrent thrombotic events in TAPS, comparing it to Support Vector Machine, Decision Tree, Gaussian Naive Bayes, and K-Nearest Neighbors. Using demographic and clinical data, model performance was assessed through multiple metrics, including accuracy, recall, specificity, precision, Youden’s Index (DYI), F1 score, Matthews Correlation Coefficient (MCC), and the area under the receiver operating characteristic curve (AUC-ROC). XGBoost outperformed all other models, achieving an AUC-ROC of 0.91, an F1-score of 91.24, and an MCC of 80.98. Recall and accuracy exceeded 92.23% and 91.35%, respectively, demonstrating robust predictive capabilities. Key predictors identified included renal insufficiency, age, and lupus anticoagulants, reinforcing the clinical relevance of these factors in risk assessment. These findings highlight the potential of XGBoost to improve risk stratification and support clinical decision-making in TAPS. By identifying critical predictors, this approach may optimize anticoagulation strategies and enhance resource allocation. However, further validation in larger cohorts and prospective studies is necessary before clinical integration.

## Introduction

Antiphospholipid Syndrome (APS) is an autoimmune disorder characterized by recurrent thrombotic events and obstetric morbidity caused by antiphospholipid antibodies (aPL), including lupus anticoagulant (LAC), anticardiolipin (aCL), and anti-β2 glycoprotein I antibodies (anti-β2GPI) ^1^. APS is a heterogeneous condition with a wide spectrum of clinical manifestations, ranging from isolated thrombotic events to obstetric complications and, in severe cases, catastrophic presentations involving multi-organ failure ^2^. The syndrome is classified into thrombotic APS (TAPS), characterized by venous, arterial, or microvascular thrombosis, and obstetric APS, involving complications such as recurrent fetal loss and placental insufficiency ^1^. APS can occur as a primary condition or in the setting of autoimmune diseases like systemic lupus erythematosus (SLE) ^3^. Thrombotic events in APS predominantly affect venous territories, with deep vein thrombosis of the lower limbs being the most common clinical presentation ^4^. Pulmonary embolism, hepatic vein thrombosis, and ischemic strokes are also frequently observed ^5^. The risk of recurrence is strongly influenced by the presence of aPL, being LAC the most pathogenic, followed by aCL and anti-β2GPI antibodies ^6–8^. Patients with triple positivity for these antibodies have the highest risk of thrombosis ^9–12^. Despite anticoagulant therapy, recurrence rates of thrombosis in patients with APS remain high. A study published in 2002 found that patients undergoing high-intensity anticoagulation therapy with an international normalized ratio (INR) target of 3.5 still experienced a recurrence rate of 9.1% patient-years, highlighting the persistent challenge of thrombotic events ^13^. Later, in 2017, another study reported that recurrence rates in APS patients treated with anticoagulants reached 23.7%, compared to 37.2% in those on antiplatelet therapy and 6.9% in those receiving combined therapy, with an estimated 20% recurrence occurring within 3.4, 7.3, and 16.3 years, respectively ^14^. More recently, in 2018, a meta-analysis of APS patients treated with direct oral anticoagulants (DOACs) reported a recurrence rate of 16%, with a mean duration of 12.5 months until the next thrombotic event, underscoring the limitations of DOACs compared to traditional anticoagulants ^15^. Given these challenges, it is urgently needed to find new methodologies to achieve the most accurate possible prediction of thrombosis recurrence in this patient population. However, traditional statistical methods for predicting thrombotic recurrence in APS are limited by their inability to capture the multifactorial and non-linear nature of the syndrome ^16^. The heterogeneity of APS, including variations in clinical presentation, antibody profiles, and response to therapy, highlights the inadequacy of conventional models for stratifying risk. Machine learning (ML) algorithms have emerged as transformative tools in medicine, offering predictive capabilities that surpass traditional methods by integrating complex, multidimensional datasets, including clinical, demographic, and laboratory variables ^17^. Unlike conventional approaches, ML models are very promising for identifying intricate patterns and interactions, making them particularly well-suited for risk prediction in APS ^18^. By addressing the limitations of conventional analysis, this study aims to assess whether ML algorithms can provide a robust framework for personalized thrombosis risk stratification in individuals with TAPS undergoing anticoagulation therapy. Among these algorithms, Extreme Gradient Boosting (XGBoost) has proven highly effective in integrating clinical data, identifying complex relationships, and improving predictive accuracy compared to traditional methods. XGBoost has been successfully applied across various medical domains, including cardiovascular event prediction ^19^, early cancer detection ^20^, and risk stratification in autoimmune diseases ^21^.

## Material and Methods

### Study Design

This retrospective study includes 72 patients with TAPS who were undergoing anticoagulation treatment and were followed at the Thrombosis and Hemostasis Unit of Hospital General Universitario Dr. Balmis in Alicante, Spain.

### Inclusion and Exclusion Criteria

Patients aged 18 years or older with a diagnosis of TAPS, confirmed by clinical thrombotic events and persistent positive aPL in at least two samples taken 12 weeks apart, and who were receiving antithrombotic treatment, were eligible for the study. Patients with incomplete clinical data were excluded. Additional exclusion criteria comprised the use of intensive immunosuppressive therapies that could interfere with the interpretation of thrombotic recurrence, documented poor treatment adherence, discontinuation of anticoagulant therapy during the follow-up period, end-stage renal disease requiring dialysis, and active cancer.

### Clinical and Laboratory Data

Variables were selected based on their known relevance to thrombotic risk in patients with TAPS. Data was extracted from an anonymized institutional database and included clinical, laboratory, and treatment-related variables documented during routine follow-up. Demographic characteristics (age and sex), common cardiovascular risk factors (including hypertension, diabetes mellitus, dyslipidemia, renal insufficiency, obesity, and smoking status), and pre-existing autoimmune conditions (such as SLE, inflammatory bowel disease, and rheumatoid arthritis) were recorded. Thrombosis-related variables encompassed the location of the first thrombotic event (venous, arterial, or small vessel) and thrombotic recurrence (the variable we aim to predict) during follow-up. Laboratory parameters included the titers of LAC, aCL (IgG and IgM), anti-β2GPI (IgG and IgM), and D-dimer levels. LAC was confirmed by dilute Russelĺs viper venom time (DRVVT) and the use of silica as a reagent sensitive to LAC derived from APTT (Diagnostica Stago, Paris, France) following manufactureŕs protocols. APL were analyzed by chemiluminescence (Werfen, Boston, USA) and D-dimer levels were determined by immunoprecipitation (Diagnostica Stago, Paris, France), in agreement with manufactureŕs instructions. Antithrombotic treatments were categorized into vitamin K antagonists (e.g., acenocoumarin, warfarin), low-molecular-weight heparin, DOACs, and antiplatelet agents. For patients on vitamin K antagonists, the last value of the International Normalized Ratio (INR) was recorded and dichotomized as 1 if INR > 60% of the upper limit of the normal range, otherwise 0.

### Predictive Machine Learning Modeling and Data Analysis

In this study, we propose the use of the XGBoost model for predicting recurrent thrombotic events and compare its performance with other ML algorithms. To evaluate the effectiveness of the proposed XGBoost model, we compared it with four ML algorithms: Support Vector Machine (SVM), Decision Tree (DT), Gaussian Naive Bayes (GNB), and K-Nearest Neighbors (KNN). Continuous variables were standardized using z-scores, and features with high collinearity (r > 0.8) were excluded to avoid redundancy. Model performance was assessed using 5-fold cross-validation with principal metrics including the area under the receiver operating characteristic curve (AUC-ROC), F1-score (the harmonic mean of precision and recall), and MCC (Matthews Correlation Coefficient), as they provide a comprehensive assessment of predictive performance. AUC-ROC evaluates the model’s ability to differentiate between classes across various thresholds, making it particularly suitable for imbalanced data. F1-score balances precision and recall, offering a reliable metric for minimizing false positives and false negatives, while MCC considers all elements of the confusion matrix, providing a robust evaluation, especially in cases of class imbalance. Recall, accuracy, specificity, precision and DYI (Youden’s Index) were used as secondary metrics, as they can be influenced by class imbalance and may introduce bias. Metrics such as recall and precision, for instance, focus on specific aspects of model performance and may not reflect overall quality, while accuracy and specificity may overestimate performance in datasets where one class dominates. On the other hand, DYI adds value as a secondary metric to identify the most predictive model by balancing the trade-offs between sensitivity and specificity. While it may not provide as comprehensive an assessment as AUC-ROC, F1-score, or MCC, it complements these metrics by offering an additional perspective on model performance. By prioritizing MCC, F1-score, and AUC-ROC, we ensured a more balanced and reliable evaluation of model efficiency. This combination enabled a thorough and objective model comparison. Hyperparameter tuning was performed using grid search optimization to identify the best model configurations. All analyses were conducted using Python (version 3.9), leveraging libraries such as Scikit-learn for model implementation, Pandas and NumPy for preprocessing, and Matplotlib for visualization. To evaluate the association between categorical variables (e.g., presence of venous location [VL], thrombophilia, renal insufficiency [RI]) and thrombotic recurrence, contingency tables were created, and the Chi-squared test of independence was used. For small sample sizes or when assumptions of the Chi-squared test were not met, Fisher’s exact test was applied. A p-value < 0.05 was considered statistically significant. All analyses were performed using Python, or SPSS, with appropriate statistical libraries.

## Results

All data used for the analysis in this study are available in supplemental material file Table S1, which includes the complete clinical characteristics of the patients. A total of 72 patients met the inclusion criteria, with a mean age of 60.5 ± 14.7 years, of whom 40% were male. Hypertension was present in 54.2% of patients, diabetes mellitus in 19.4%, and renal insufficiency in 15.3%. Laboratory results showed that 30.6% of patients were positive for LAC, 4.2% for aCL antibodies, and 26.4% for anti-β2GPI. Triple positivity was not observed. A comparison of clinical and demographic variables in patients with and without thrombosis recurrence is summarized in Table 1.

**Table 1.**
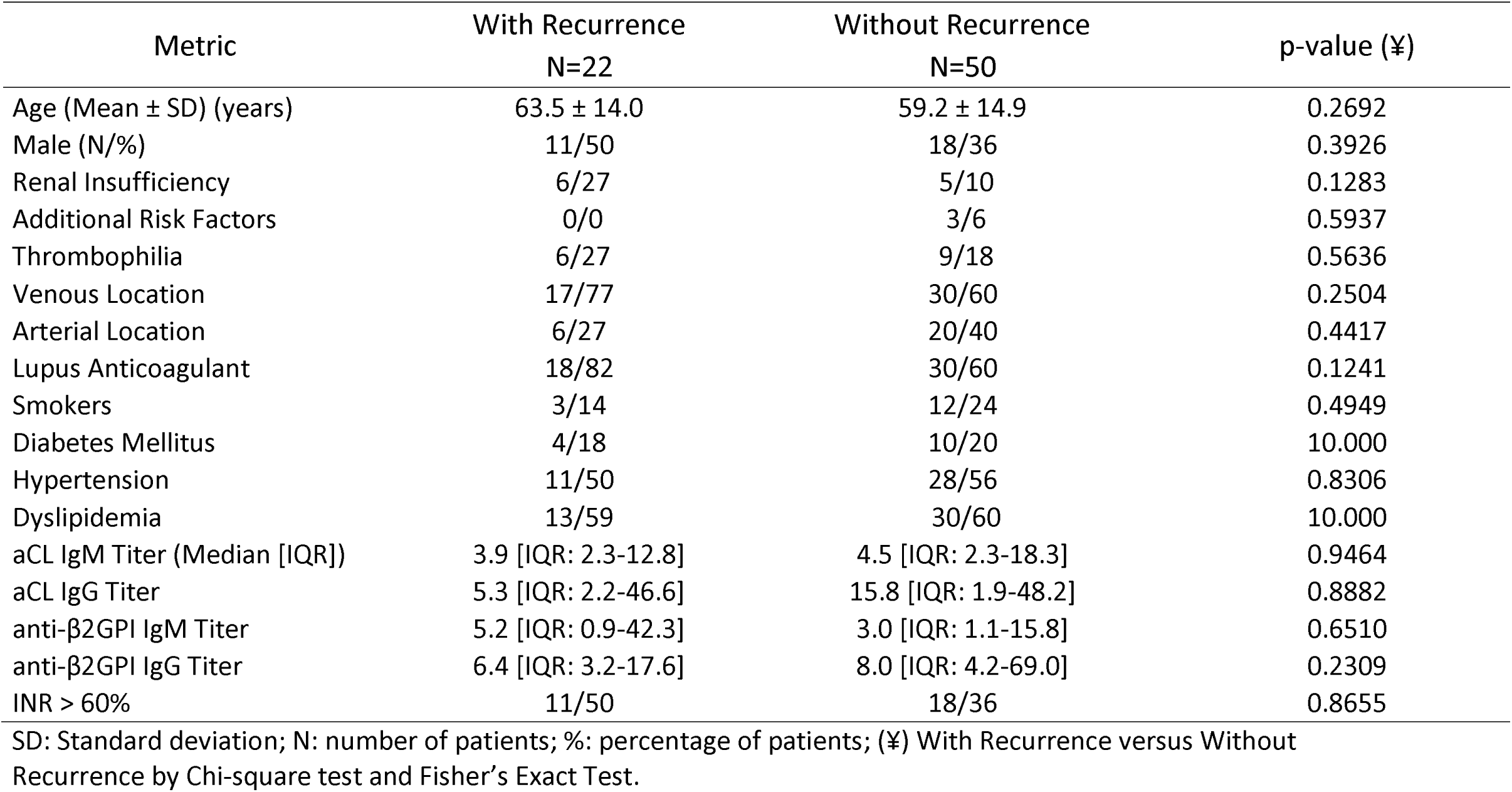
Comparison of clinical and demographic variables in patients with and without thrombosis recurrence.

A comprehensive comparison of model performance metrics is presented in Table 2. Among the ML algorithms evaluated, XGBoost demonstrated superior performance during the testing phase. The relative contributions of the study variables are illustrated in Figure 1. The feature importance analysis of XGB identified renal insufficiency, LAC, VL, and age as the most significant predictors of thrombotic recurrence. XGBoost achieved an AUC-ROC of 0.9125, an F1-score of 91.24, and an MCC of 80.98, with recall and accuracy exceeding 92.23% and 91.35%, respectively, highlighting its robust predictive capability compared to other models Figures 2-4.

**Table 2.**
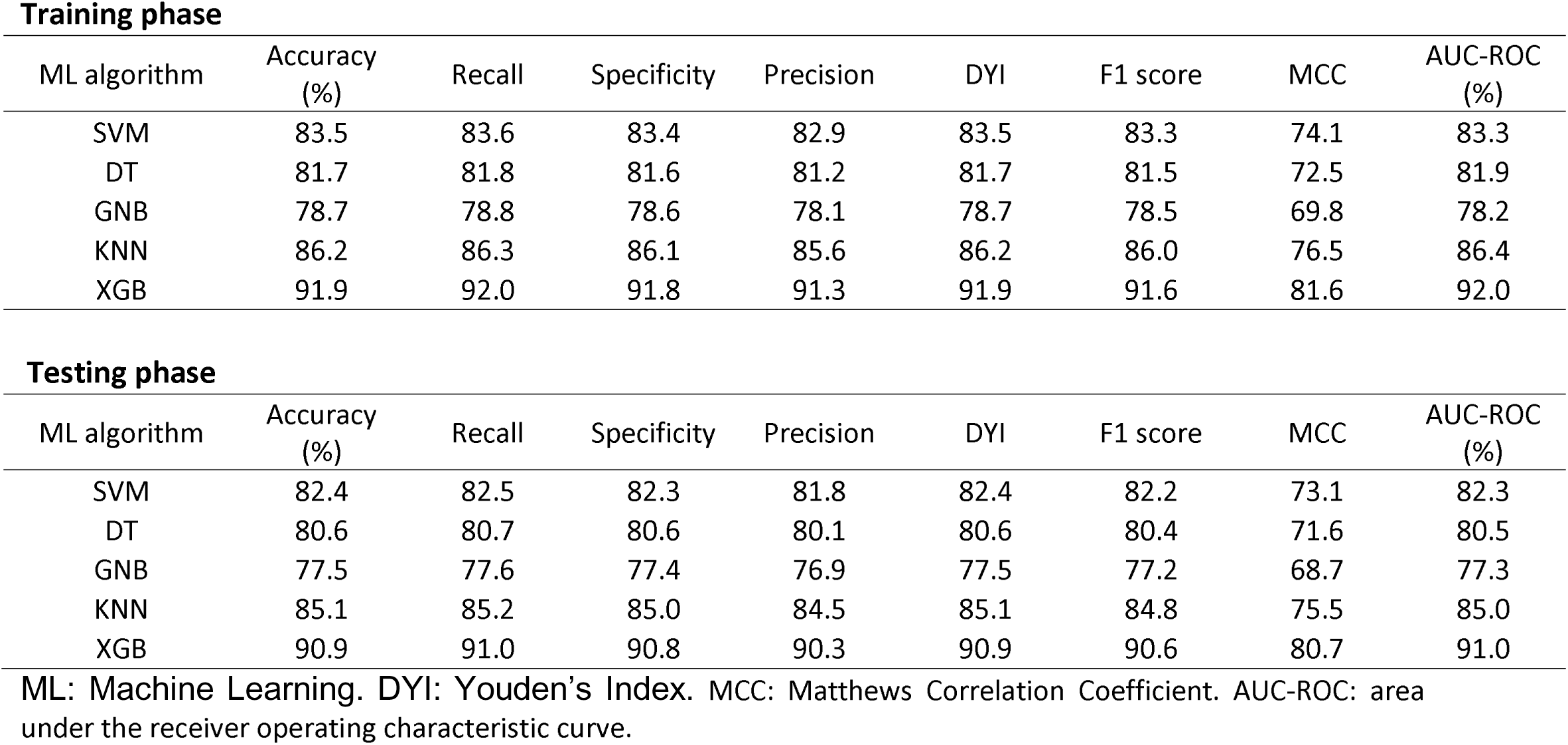
Performance metrics achieved during the training and testing phases of the machine learning models.

| **Training phase** |  |  |  |  |  |  |  |  |
| --- | --- | --- | --- | --- | --- | --- | --- | --- |
| ML algorithm | Accuracy (%) | Recall | Specificity | Precision | DYI | F1 score | MCC | AUC-ROC (%) |
| SVM | 83.5 | 83.6 | 83.4 | 82.9 | 83.5 | 83.3 | 74.1 | 83.3 |
| DT | 81.7 | 81.8 | 81.6 | 81.2 | 81.7 | 81.5 | 72.5 | 81.9 |
| GNB | 78.7 | 78.8 | 78.6 | 78.1 | 78.7 | 78.5 | 69.8 | 78.2 |
| KNN | 86.2 | 86.3 | 86.1 | 85.6 | 86.2 | 86.0 | 76.5 | 86.4 |
| XGB | 91.9 | 92.0 | 91.8 | 91.3 | 91.9 | 91.6 | 81.6 | 92.0 |
| **Testing phase** |  |  |  |  |  |  |  |  |
| ML algorithm | Accuracy (%) | Recall | Specificity | Precision | DYI | F1 score | MCC | AUC-ROC (%) |
| SVM | 82.4 | 82.5 | 82.3 | 81.8 | 82.4 | 82.2 | 73.1 | 82.3 |
| DT | 80.6 | 80.7 | 80.6 | 80.1 | 80.6 | 80.4 | 71.6 | 80.5 |
| GNB | 77.5 | 77.6 | 77.4 | 76.9 | 77.5 | 77.2 | 68.7 | 77.3 |
| KNN | 85.1 | 85.2 | 85.0 | 84.5 | 85.1 | 84.8 | 75.5 | 85.0 |
| XGB | 90.9 | 91.0 | 90.8 | 90.3 | 90.9 | 90.6 | 80.7 | 91.0 |

**ML: Machine Learning. DYI: Youden's Index. MCC: Matthews Correlation Coefficient. AUC-ROC: area under the receiver operating characteristic curve.**

**Figure 1.**
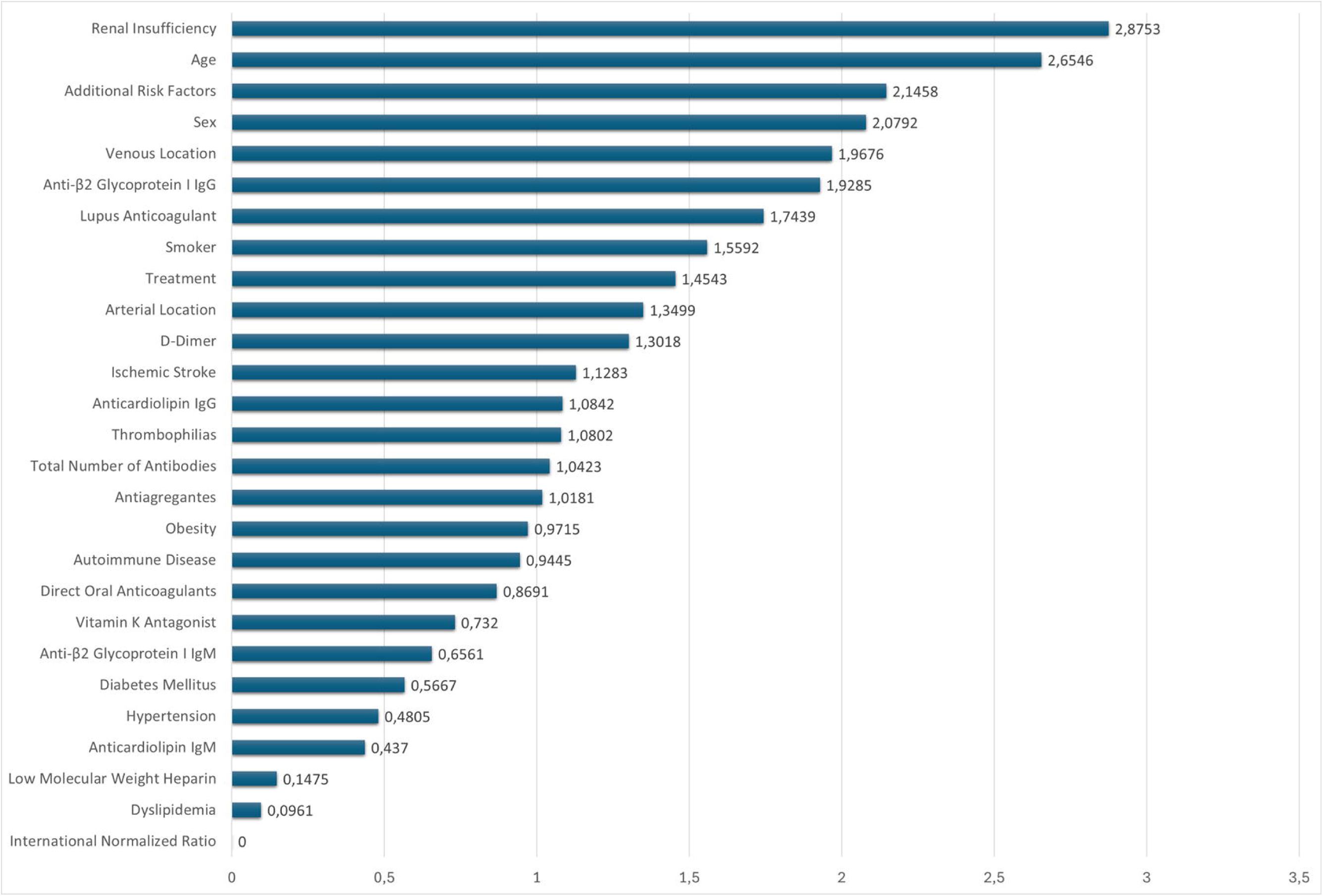
Variable and weights assigned to each variable, indicating their relative contribution to the model.

**Figure 2.**
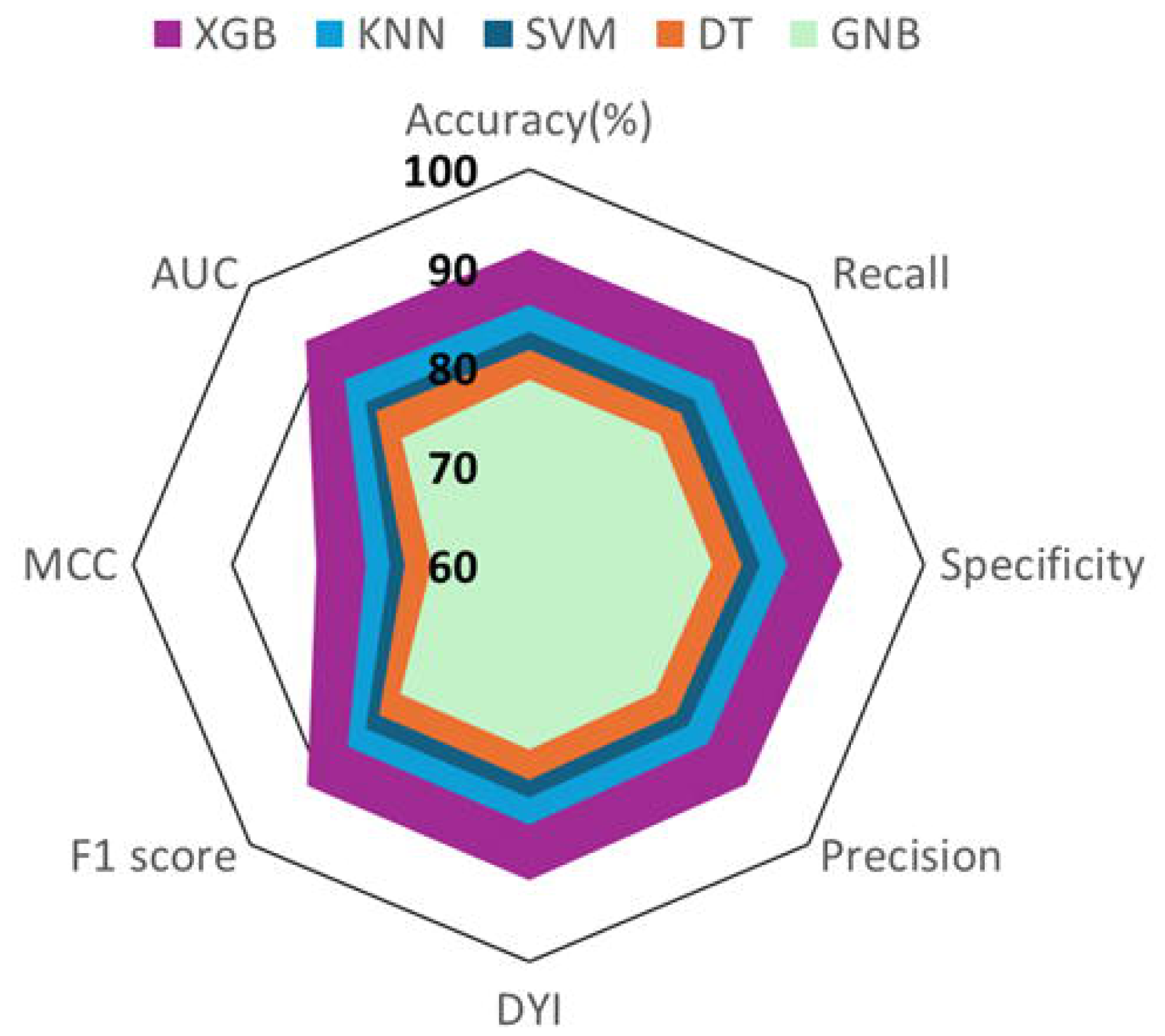
Radar Chart of machine learning algorithm performance during the training phase. XGB: Extreme Gradient Boosting. KNN: K-nearest neighbors. SVM: Support Vector Machine. DT: Decision Tree. GNB: Gaussian Naive Bayes

**Figure 3.**
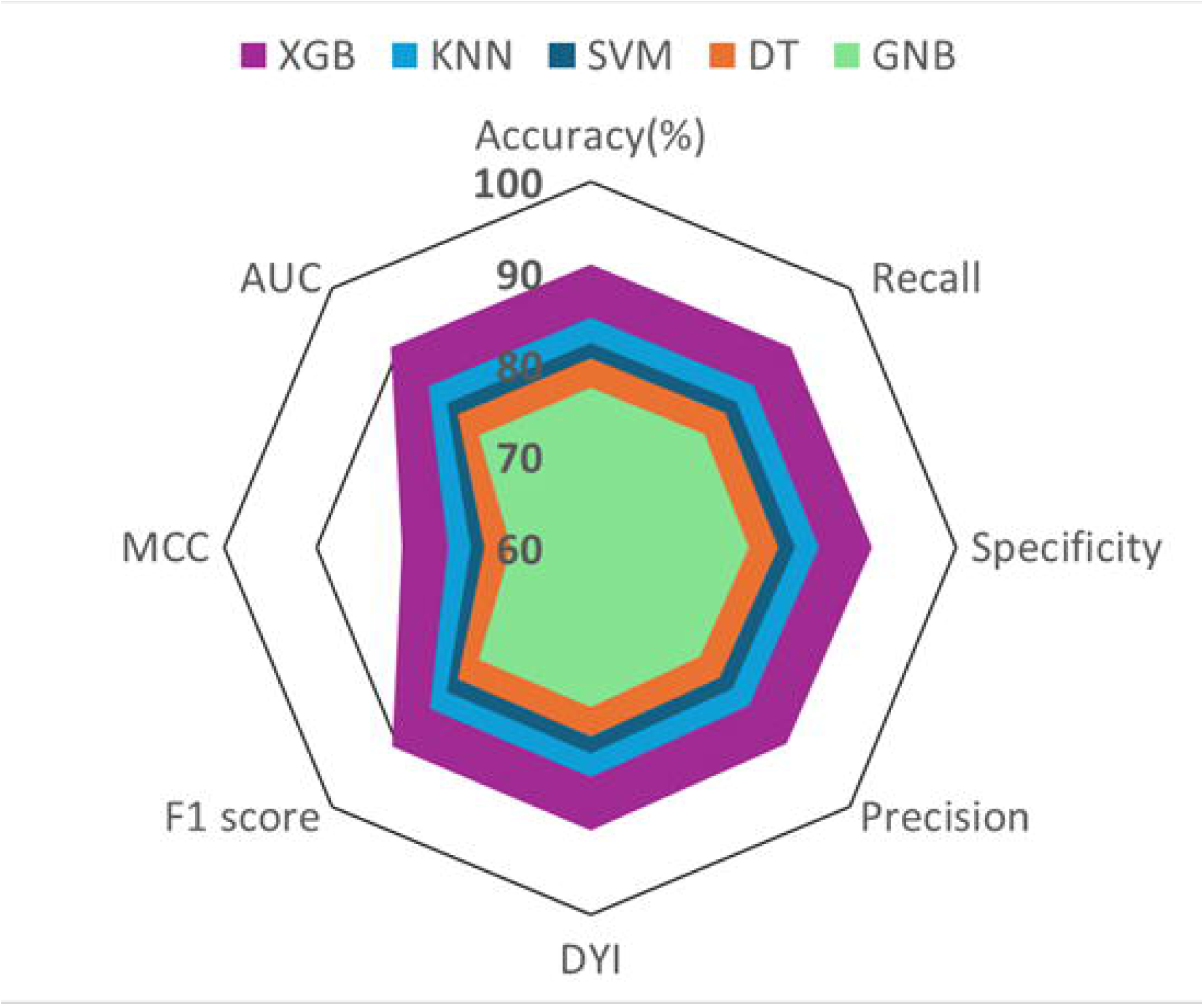
Radar Chart of machine learning algorithm performance during the testing phase. XGB: Extreme Gradient Boosting. KNN: K-nearest neighbors. SVM: Support Vector Machine. DT: Decision Tree. GNB: Gaussian Naive Bayes

**Figure 4.**
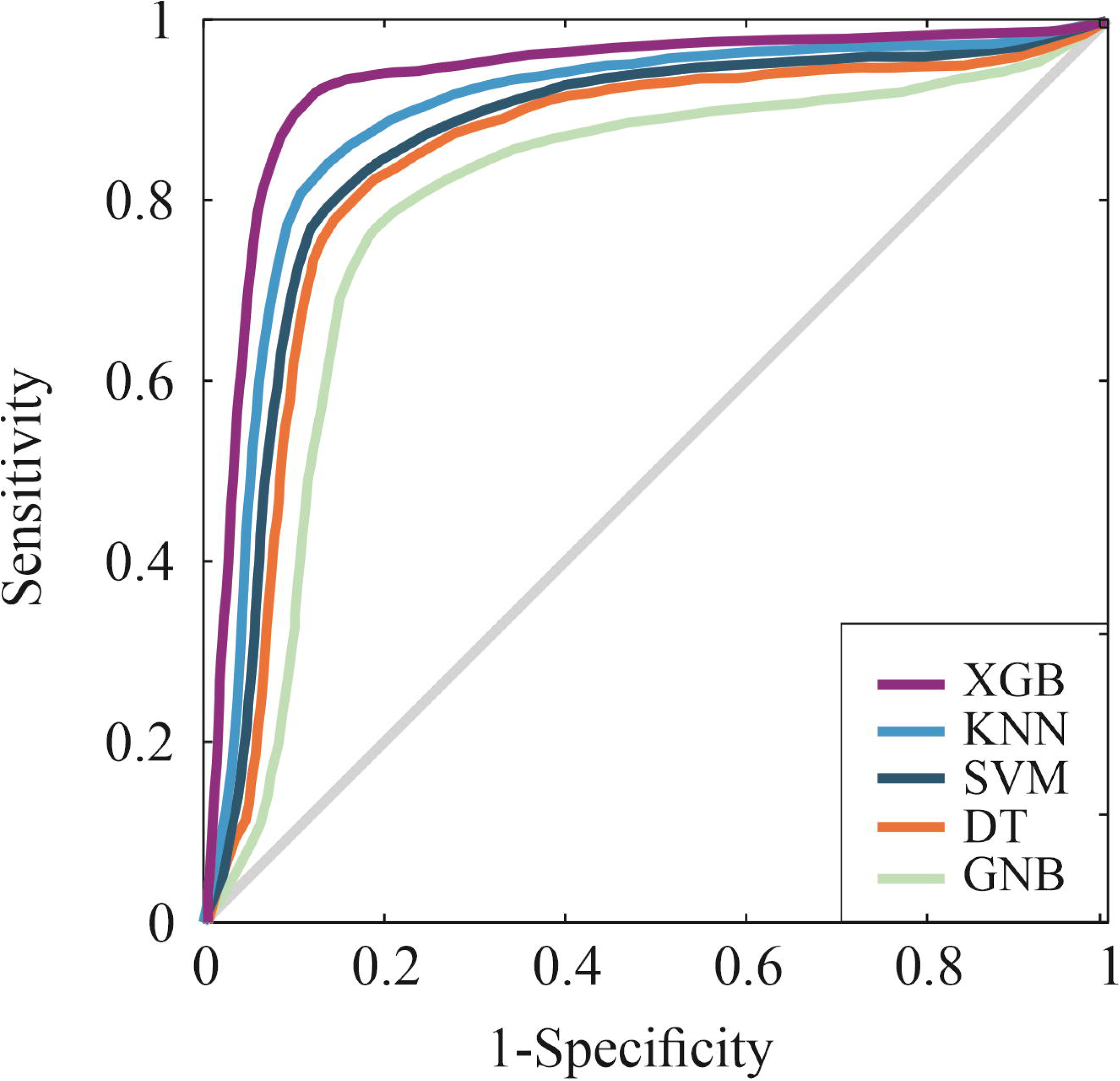
A multi-model ROC curve comparing the AUC-ROC of all five algorithms. XGB: Extreme Gradient Boosting. KNN: K-nearest neighbors. SVM: Support Vector Machine. DT: Decision Tree. GNB: Gaussian Naive Bayes

## Discussion

The findings of this study underscore the potential of ML, particularly XGBoost, in predicting recurrent thrombotic events in patients with TAPS. XGBoost outperformed traditional predictive models by effectively integrating clinical, demographic, and laboratory variables to generate accurate risk assessments. Its ability to handle heterogeneous data, detect complex interactions among multiple variables, and mitigate overfitting through regularization contributes to its superior predictive capabilities also observed in previous studies ^22–24^.

The identification of renal insufficiency, age, and LAC as key predictors aligns with existing clinical knowledge, reinforcing the understanding that renal dysfunction exacerbates endothelial damage and significantly contributes to thrombotic risk ^25^. Age, a well-established risk factor, is associated with physiological changes such as arterial stiffening, endothelial dysfunction, and increased coagulability, all of which elevate thrombotic risk ^26^. Additionally, aging is linked to enhanced activation of the coagulation cascade and elevated levels of pro-inflammatory biomarkers that promote thrombosis ^27^. Older patients frequently present with comorbidities, including hypertension, diabetes, and dyslipidemia, which further contribute to vascular dysfunction and increase the likelihood of recurrent thrombotic events ^28^. The presence of LAC has been identified as the most potent thrombotic risk factor in APS, significantly increasing the risk of arterial and venous thrombosis ^12^. The presence of LAC in patients with TAPS has been associated with a significant increase in resistance to activated protein C (APC), a key mechanism in coagulation regulation ^29^. This resistance decreases APC’s ability to inactivate factors Va and VIIIa, promoting a prothrombotic state due to higher levels of thrombin generation after coagulation activation. Excess thrombin subsequently activates thrombin-activatable fibrinolysis inhibitor, an enzyme that stabilizes the clot by removing plasminogen-binding sites on fibrin, thereby reducing plasminogen conversion into plasmin and blocking fibrinolysis ^30^. In addition, enhanced thrombin generation also induces the production of plasminogen activator inhibitor-1 (PAI-1) ^31^, a potent fibrinolysis suppressor that prevents the activation of plasminogen by tissue plasminogen activator (t-PA) ^32^. Elevated PAI-1 levels further contribute to fibrin accumulation and persistent clot formation. Together, these mechanisms generate a prothrombotic state in TAPS patients, explaining the role of LAC as a key risk factor for arterial and venous thrombosis in this population. In addition to the presence of LAC, the presence of aCL and anti-β2GPI antibodies are contributing variables to our predictive model, which aligns with previous findings indicating that triple positivity has been strongly associated with a higher likelihood of recurrent thrombotic events in TAPS patients ^33^.

A significant observation in this study is that the INR did not exhibit predictive value for thrombotic recurrence. This may be attributed to the presence of confounding factors not captured by the model, and/or that INR alone is not a reliable predictor of thrombotic events ^34–36^. While INR is a standard tool for monitoring anticoagulation with vitamin K antagonists, recent studies suggest that it does not always accurately reflect the procoagulant state in APS patients, which may explain its limited utility in our predictive model ^37,38^.

### Limitations

Although the sample size in this study may be considered a limitation, recent evidence suggests that ML models can achieve reliable performance even in small cohorts when appropriate validation techniques, such as cross-validation, are applied ^39^. To further mitigate the impact of the limited sample size, this study employed data synthesis techniques, a strategy proven to enhance the performance of ML models in prior studies ^40^. To improve predictive performance, future research should consider incorporating additional aPL, such as anti-phosphatidylserine/prothrombin and anti-annexin-V, which have demonstrated strong associations with thrombotic events in recent studies ^41,42^. Additionally, platelet lipidomic may offer valuable insights into thrombotic risk stratification ^43,44^. In addition, altogether using a larger, multicenter cohorts design could further improve the accuracy and applicability of the predictive models. From a clinical perspective, the adoption of ML tools like XGBoost has the potential to revolutionize patient care by enabling personalized risk stratification and optimizing anticoagulation strategies. However, for successful real-world implementation, user-friendly interfaces and integration into electronic health record systems will be essential to facilitate clinical decision-making.

### Conclusions

This study demonstrates the potential of XGBoost in TAPS management, achieving an AUC-ROC of 0.91, an F1-score of 91.24, and an MCC of 80.98, with recall and accuracy exceeding 92.23% and 91.35%, respectively, highlighting its robust predictive capability compared to other models. Key predictors such as renal insufficiency, age, and LAC highlight the importance of integrating clinical and laboratory data for precise risk stratification. Integrating multi-omics data, such as platelet lipidomic, and additional aPL, like anti-phosphatidylserine/prothrombin and anti-annexin-V, could boost predictive accuracy by uncovering molecular interactions and pathways driving thrombotic risk in platelets, the main target of aPL antibodies. These approaches provide a comprehensive view of the biological processes underlying TAPS, enabling the identification of novel biomarkers and therapeutic targets, ultimately improving patient outcomes and resource allocation in TAPS care.

## Data Availability

All data produced in the present study are available upon reasonable request to the authors

## Abbreviations

APS: Antiphospholipid Syndrome
aPL: Antiphospholipid antibodies
LAC: Lupus anticoagulant
aCL: Anticardiolipin
anti-β2GPI: Anti-β2 glycoprotein I antibodies
TAPS: Thrombotic Antiphospholipid Syndrome
SLE: Systemic Lupus Erythematosus
INR: International Normalized Ratio
DOACs: Direct Oral Anticoagulants
ML: Machine Learning
XGBoost: Extreme Gradient Boosting
SVM: Support Vector Machine
DT: Decision Tree
GNB: Gaussian Naive Bayes
KNN: K-Nearest Neighbors
DRVVT: Dilute Russell’s viper venom time
APTT: Activated Partial Thromboplastin Time
DYI: Youden’s Index
MCC: Matthews Correlation Coefficient
AUC-ROC: Area Under the Receiver Operating Characteristic Curve
VL: Venous Location
RI: Renal Insufficiency
APC: Activated Protein C
PAI-1: Plasminogen Activator Inhibitor-1 t
PA: Tissue Plasminogen Activator

## Acknowledgements

The authors thank Bayer Hispania, S.L. for its contribution to the creation of the *Cátedra de Inteligencia Artificial*, an academic initiative focused on Artificial Intelligence-driven data analysis, which carried out the data analysis for this study.

## Author Contributions

Conceptualization, A.M.R., I.F.B. and P.M.V.; Methodology, A.M.R., I.F.B. and P.M.V.; Software, J.M.S.; Formal analysis, J.M.S.; Data curation, A.M.R., I.F.B., J.M.S and P.M.V.; Writing – original draft preparation, I.F.B.; Writing – review and editing, all authors; Visualization, J.M.S.; Supervision, I.F.B and P.M.V.; All authors have read and agreed to the published version of the manuscript.

## Availability of Data and Materials

The data supporting the findings of this study are provided as supplemental material in File 1.

## Consent for Publication

Written informed consent was obtained from patients or legal guardians before inclusion.

## Ethics Approval and Consent to Participate

The study was approved by the local ethical committee.

## Supplemental material

Supplemental material for this article is available online in the file Table S1.

